# “I feel under attack”: Transgender Adolescents and Caregivers Opposition to Restrictions on Gender-Affirming Medical Care in Brazil

**DOI:** 10.1101/2025.09.10.25335522

**Authors:** Bruna Caruso Mazzolani, Luana Cordeiro de Oliveira, Igor Longobardi, Fabiana Infante Smaira, Fernanda Sabatini, Fernanda Evangelista Bandeira de Melo, Samuel Bittar, Thifany Helena Torres, Fernanda Baeza Scagliusi, Alexandre Saadeh, Hamilton Roschel, Bruno Gualano

## Abstract

In April 2025, Brazil’s Federal Council of Medicine (CFM) issued Resolution 2.427/2025 banning puberty blockers, restricting hormone therapy until age 18, and delaying gender-affirming surgeries until age 21. These measures sharply contrast with previous national protocols aligned with international standards of care. We conducted a qualitative study to explore how transgender adolescents and their caregivers perceive these new restrictions. Nine focus groups were held between May and June 2025 with 31 transgender adolescents (11–18 years) and 21 caregivers receiving care at Brazil’s largest gender identity outpatient clinic. Discussions were transcribed verbatim, coded, and analyzed using qualitative content analysis. Participants consistently viewed the resolution as unjust, ideologically motivated, and detached from scientific evidence. Adolescents described puberty blockers and hormone therapy as life-saving, emphasizing their positive physical and psychological effects. They rejected detransition as a valid justification for restrictions, framing it as rare and often driven by external pressures. Caregivers echoed these concerns, describing the resolution as harmful, unscientific, and likely to exacerbate inequalities, mental health challenges, and unsafe self-medication practices. Both groups highlighted that gender-affirming care is preceded by long-term, multidisciplinary follow-up under robust clinical protocols, minimizing risks of regret or adverse outcomes. Findings show that restrictive policies are not perceived as neutral regulation but as an assault on the rights, health, and dignity of transgender youth. By amplifying the lived experiences of adolescents and caregivers, this study points out the urgent need for evidence-based, inclusive, and participatory health policies that do not exclude trans voices.

## INTRODUCTION

In recent years, gender-affirming care for transgender and gender-diverse adolescents has been increasingly threatened by restrictive policies across different countries. In the UK and USA, laws have been enacted to ban or limit access to puberty blockers, hormone therapy, and gender-affirming surgeries (House, 2025; NHSE, 2024), exacerbating a globally recognized issue in which healthcare systems for transgender individuals often exhibit insufficient or even negligible investment in resources (Coleman et al., 2022).

A similar trend has recently emerged in Brazil. In April 2025, the Federal Council of Medicine (CFM) - the national regulatory authority for physicians - issued Resolution 2.427/2025 (Medicina, 2025) banning puberty blockers, prohibiting hormone therapy before age 18, restricting some surgical procedures until age 21, and imposing additional constraints on clinical care for transgender individuals. These restrictions represent a sharp departure from Brazil’s prior clinical practice, previously aligned with international standards (Coleman et al., 2022; Hembree et al., 2017; Oliphant et al., 2018; Rafferty, 2018; Telfer et al., 2018).

The resolution has generated widespread concern among health professionals, researchers, and advocacy groups, who argue that changes are not grounded in scientific evidence and fail to consider the lived experiences of transgender individuals (Longobardi et al., 2025). Although research gaps in the area exist, gender-affirming interventions are founded on decades of clinical experience and research, and therefore are not regarded as experimental, cosmetic, or merely a matter of patient convenience (Coleman et al., 2022). Available evidence shows that gender-affirming medical care is associated with improved mental health outcomes, reduced gender dysphoria and suicidality, and enhanced quality of life among transgender adolescents (Costa et al., 2015; de Vries et al., 2014; de Vries et al., 2011; Kuper et al., 2020; Kristina R. Olson, 2025; Tordoff et al., 2022), while treatment regret or discontinuation remains low (Boskey et al., 2025; Cavve et al., 2024; K. R. Olson et al., 2024; van der Loos et al., 2022). Conversely, restricting access to such care may further entrench the structural and psychosocial vulnerabilities experienced by this population.

In Brazil, transgender individuals face severe systemic barriers to public healthcare, including the geographic concentration of services, scarcity of specialized outpatient care and professionals, long waiting times, and frequent disregard for identity and chosen names (Bender et al., 2025; Pfeil & Alves, 2024; Rocon et al., 2019). These challenges often lead to treatment abandonment, misguided self-care practices, or unsafe self-medication, such as unsupervised hormone or industrial silicone use (Bender et al., 2025; Pfeil & Alves, 2024; Rocon et al., 2019; Velasco et al., 2022). State neglect (Benevides, 2023; Pfeil & Alves, 2024) and health insurance denials (Transmasculinidades, 2023) frequently force individuals to seek court orders for access to hormone therapy and surgeries. Such disparities and institutionalized violence contribute to anxiety, depression, poorer quality of life, and high rates of self-harm and suicide (Barroso et al., 2025; Benevides, 2023; Hunter et al., 2021; Pfeil & Alves, 2024; Transmasculinidades, 2023). In fact, gender-diverse adolescents, particularly ages 16–25, report elevated anxiety, depression, and reduced well-being due to minority stress from discrimination and stigma (Hunter et al., 2021; Velasco et al., 2022). Between 2018 and 2021, transgender youth accounted for over half of 2,761 self-inflicted injury reports among transgender women and *travestis* in Brazil (Alves & Semente, 2023; Benevides, 2023). Therefore, institutionalized violence and transphobia create a hostile social environment that profoundly affects the mental health and safety of transgender adolescents (Benevides, 2023; Pfeil & Alves, 2024; Transmasculinidades, 2023). Against this backdrop of systemic inequities, recent policy debates have further intensified, yet the perspectives of those directly affected (i.e., transgender adolescents and their caregivers) remain underrepresented.

In a recent survey conducted at Brazil’s largest outpatient gender identity clinic, an overwhelming majority of both adolescents and caregivers expressed strong opposition to the new resolution (unpublished data). However, quantitative data alone cannot fully capture the complexity of the lived experiences, emotional responses, and actions taken by those impacted. As emphasized by participatory models of health policy design (Boenink et al., 2018), understanding the subjective experiences of patients is critical to building equitable, intersectional, effective, and person-centered care systems.

As Sandelowski (2000)(Sandelowski, 2000) suggests, qualitative descriptive research can provide a rich, detailed description of a particular group’s perspectives or understandings regarding a phenomenon about which little is currently known. Therefore, this study aimed to qualitatively explore the opinions and perceptions of transgender adolescents and transgender children and adolescents’ caregivers regarding the new CFM resolution. By centering these voices, the study seeks to inform more ethical, evidence-based, and inclusive policies for transgender healthcare.

## METHODS

This manuscript presents an analysis of a larger study, using a descriptive qualitative approach with focus group discussions to explore the experiences of transgender adolescents and their caregivers regarding lifestyle. With the promulgation of the new CFM resolution coinciding with the commencement of the study, we adapted the focus group script to capture participants’ perceptions on the new resolution. Topics related to gender variability, interpersonal relationships, future expectations, perceptions and knowledge about lifestyle, relationship with the body, and opinions and perceptions related to the new CFM resolution were explored. Herein, we focused on analyzing the content related to the new CFM resolution.

The study was approved by the local Ethical Committee (Commission for Analysis of Research Projects, CAPPesq; approval: 83553224.2.0000.0068). Participants provided signed informed assent (those under 18 years) and/or consent before participation.

### Participants

Eligible participants were receiving care at the Transdisciplinary Gender Identity and Sexual Orientation Service at the Clinical Hospital of the University of São Paulo Medical School at the time of the study. Transgender adolescents (aged 11–18 years) and their caregivers, as well as caregivers of transgender children (aged under 11) were recruited in person during clinical visits or remotely via phone, messaging apps and email with prior authorization. Participants were provided with explanations of the qualitative data collection design and purpose. Descriptive sociodemographic and clinical data, including mental health conditions, were obtained from medical records.

### Study design

We conducted nine focus group meetings (four with transgender adolescents and five with caregivers) at Brazil’s largest outpatient gender identity service between May and June 2025. Focus groups were held separately for adolescents and caregivers. Caregiver participation occurred regardless of whether their children or adolescents attended, whereas adolescent participation was independent of the involvement of their respective caregivers. Participants were invited from a pre-established list of those scheduled for clinical visits on focus group days and randomly selected to ensure group composition. For each session, 10–15 adolescents were invited to obtain 7–10 participants, and 6–8 caregivers were invited to yield 4–8 participants, with a higher target for adolescents given the expected challenges in engagement in the discussion at this age.

The discussions occurred in a quiet, private room and lasted between 60 and 100 minutes. Each meeting began with an explanation of the purpose and procedures of the discussion. Oral consent to record the discussion was obtained with assurance that audiotape’s transcription would be in strict confidentiality and ensuring anonymity. A moderator conducted the discussion following a script of open questions previously structured (SM1 in Supplementary material). A pilot focus group was conducted to trial the script and changes were made to improve facilitated conversation, to refine flow and structure of questions, and to ensure clarity of communication. Data obtained in this pilot group were not considered for subsequent analyzes. One experienced qualitative researcher (LCO, FS, FE or THT) moderated the focus group discussions, and one or two trained co-moderators (BCM, LCO, FIS, FS, FE, and/or THT) facilitated seating and took field notes of nonverbal cues.

### Data analysis

Quantitative data describing participants’ characteristics are presented as mean ± standard deviation and range [min⎯max] for continuous variables or frequency and percentage for categorical variables. BCM, IL, FIS and SM transcribed focus group audio recording *verbatim* and reviewed all transcriptions and audio recordings to ensure accuracy. Given the objective of this study, the content of the following questions of the transgender adolescents’ version of the interview guide were analyzed, as well as their adapted version for caregivers: “What would it be like to be able to block puberty, or what is it like to be able to block puberty? And what is it like to think about hormone therapy and surgeries? Do you feel like you need to go through them? What do you hope will happen if you go through them?”, “Have you heard about the new resolution from the Federal Council of Medicine? How do you feel about the news that puberty blocking may no longer be possible? What do you think of the justification for this decision?”, “And how do you feel about the news that the minimum age for hormone therapy may become 18 and for some surgeries 21? What do you think of the justification for this decision?” and “What have you and your guardians been saying about this news? What have you done or would like to do about it?” (SM1 in Supplementary Material). Additionally, the words “resolution”, “law”, “Federal Council of Medicine” and “CFM” were located all over the transcripts to ensure all content related to the new resolution was considered. Excerpts that had these words but with an incongruous context to our aims (e.g., talking about other laws) were excluded. Analyses were conducted in Portuguese to ensure that linguistic nuance was maintained, and quotes were translated for inclusion in the manuscript.

A content analysis was conducted, following the process described by Burnard et al. (2008)(Burnard et al., 2008), with both deductive and inductive approaches. An initial code system was developed based on the moderator’s guide and study objectives. Then, an inductive approach was performed utilizing the technique of cutting and sorting (Bernard et al., 2016), with support of the MAXQDA data management software.

The first stage of the method consisted of taking notes in the form of a memo, during the initial reading of the transcriptions, made by two researchers independently (BCM and LCO). In the next stage, “open coding” was carried out by one researcher (BCM), with the central meanings of each excerpt being highlighted, following the construction of a codebook as described by (MacQueen et al., 1998b), in addition to Burnard’s method (SM2 in Supplementary Material). Next, the codebook was discussed by two researchers (BCM and LCO) and the results were validated by the research team. Two researchers (BCM and LCO) coded the data independently. The kappa coefficient for interrater reliability were calculated and both categorizations were discussed together, aiming to address discordances. Final kappa coefficient was considered adequate (kappa = 0.970) (Cohen, 1960). The codebook presents for each theme a brief and extended description; inclusion and exclusion criteria; typical and atypical quotes present in the grouped statements; and an example of a citation classified as “close but no” (MacQueen et al., 1998a).

Participants’ speeches were anonymized by using the letter A followed by a number between 1 and 31 for transgender adolescents and the letter C followed by a number between 1 and 21 for caregivers.

## RESULTS

### Participants and context

A total of 52 transgender adolescents and 32 caregivers were invited to participate. Seven adolescents did not respond, eight were unavailable, and six confirmed participation but did not attend. Among caregivers, four did not respond, five were unavailable, and two confirmed participation but did not attend. Therefore, 31 transgender adolescents and 21 caregivers representing 7 children and 14 adolescents took part in the study. Characteristics of transgender adolescents and transgender children and adolescents represented by their caregivers are presented in Table 1. Table 2 presents characteristics of the caregivers. Only 10 adolescents had their respective caregivers participating in the study, while the remaining 21 adolescents and 11 caregivers participated independently.

**Table 1.**
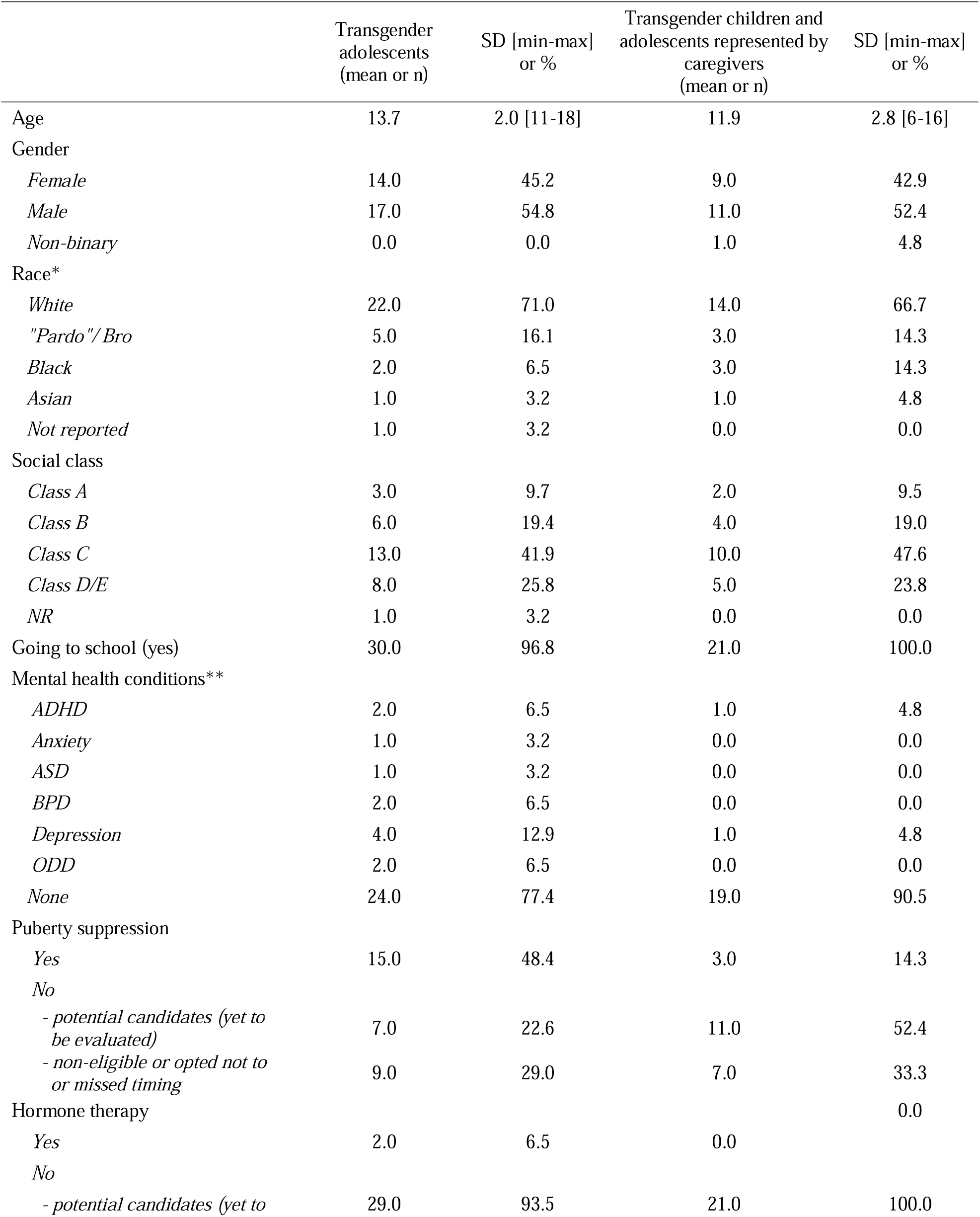
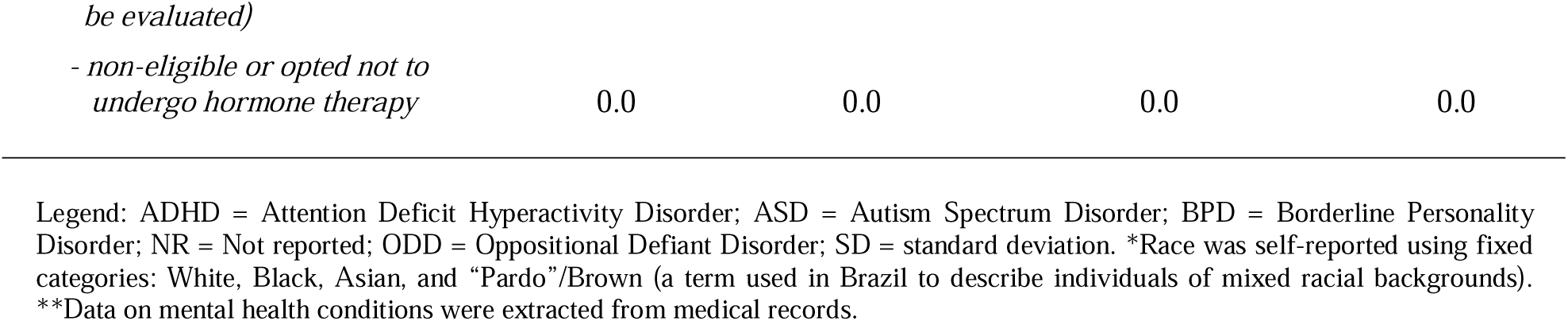
Demographic and Clinical Characteristics of Transgender Adolescents (n=31) and Transgender Children and Adolescents represented by their caregivers (n=21).

|  | Transgender adolescents<br>(mean or n) | SD [min-max]<br>or % | Transgender children and<br>adolescents represented by<br>caregivers<br>(mean or n) | SD [min-max]<br>or % |
| --- | --- | --- | --- | --- |
| Age | 13.7 | 2.0 [11-18] | 11.9 | 2.8 [6-16] |
| Gender |  |  |  |  |
| *Female* | 14.0 | 45.2 | 9.0 | 42.9 |
| *Male* | 17.0 | 54.8 | 11.0 | 52.4 |
| *Non-binary* | 0.0 | 0.0 | 1.0 | 4.8 |
| Race* |  |  |  |  |
| *White* | 22.0 | 71.0 | 14.0 | 66.7 |
| *"Pardo"/ Bro* | 5.0 | 16.1 | 3.0 | 14.3 |
| *Black* | 2.0 | 6.5 | 3.0 | 14.3 |
| *Asian* | 1.0 | 3.2 | 1.0 | 4.8 |
| *Not reported* | 1.0 | 3.2 | 0.0 | 0.0 |
| Social class |  |  |  |  |
| *Class A* | 3.0 | 9.7 | 2.0 | 9.5 |
| *Class B* | 6.0 | 19.4 | 4.0 | 19.0 |
| *Class C* | 13.0 | 41.9 | 10.0 | 47.6 |
| *Class D/E* | 8.0 | 25.8 | 5.0 | 23.8 |
| *NR* | 1.0 | 3.2 | 0.0 | 0.0 |
| Going to school (yes) | 30.0 | 96.8 | 21.0 | 100.0 |
| Mental health conditions** |  |  |  |  |
| *ADHD* | 2.0 | 6.5 | 1.0 | 4.8 |
| *Anxiety* | 1.0 | 3.2 | 0.0 | 0.0 |
| *ASD* | 1.0 | 3.2 | 0.0 | 0.0 |
| *BPD* | 2.0 | 6.5 | 0.0 | 0.0 |
| *Depression* | 4.0 | 12.9 | 1.0 | 4.8 |
| *ODD* | 2.0 | 6.5 | 0.0 | 0.0 |
| *None* | 24.0 | 77.4 | 19.0 | 90.5 |
| Puberty suppression |  |  |  |  |
| *Yes* | 15.0 | 48.4 | 3.0 | 14.3 |
| *No* |  |  |  |  |
| - *potential candidates (yet to be evaluated)* | 7.0 | 22.6 | 11.0 | 52.4 |
| - *non-eligible or opted not to or missed timing* | 9.0 | 29.0 | 7.0 | 33.3 |
| Hormone therapy |  |  |  | 0.0 |
| *Yes* | 2.0 | 6.5 | 0.0 |  |
| *No* |  |  |  |  |
| - *potential candidates (yet to* | 29.0 | 93.5 | 21.0 | 100.0 |

Legend: ADHD = Attention Deficit Hyperactivity Disorder; ASD = Autism Spectrum Disorder; BPD = Borderline Personality Disorder; NR = Not reported; ODD = Oppositional Defiant Disorder; SD = standard deviation. \*Race was self-reported using fixed categories: White, Black, Asian, and “Pardo”/Brown (a term used in Brazil to describe individuals of mixed racial backgrounds). \*\*Data on mental health conditions were extracted from medical records.

**Table 2.**
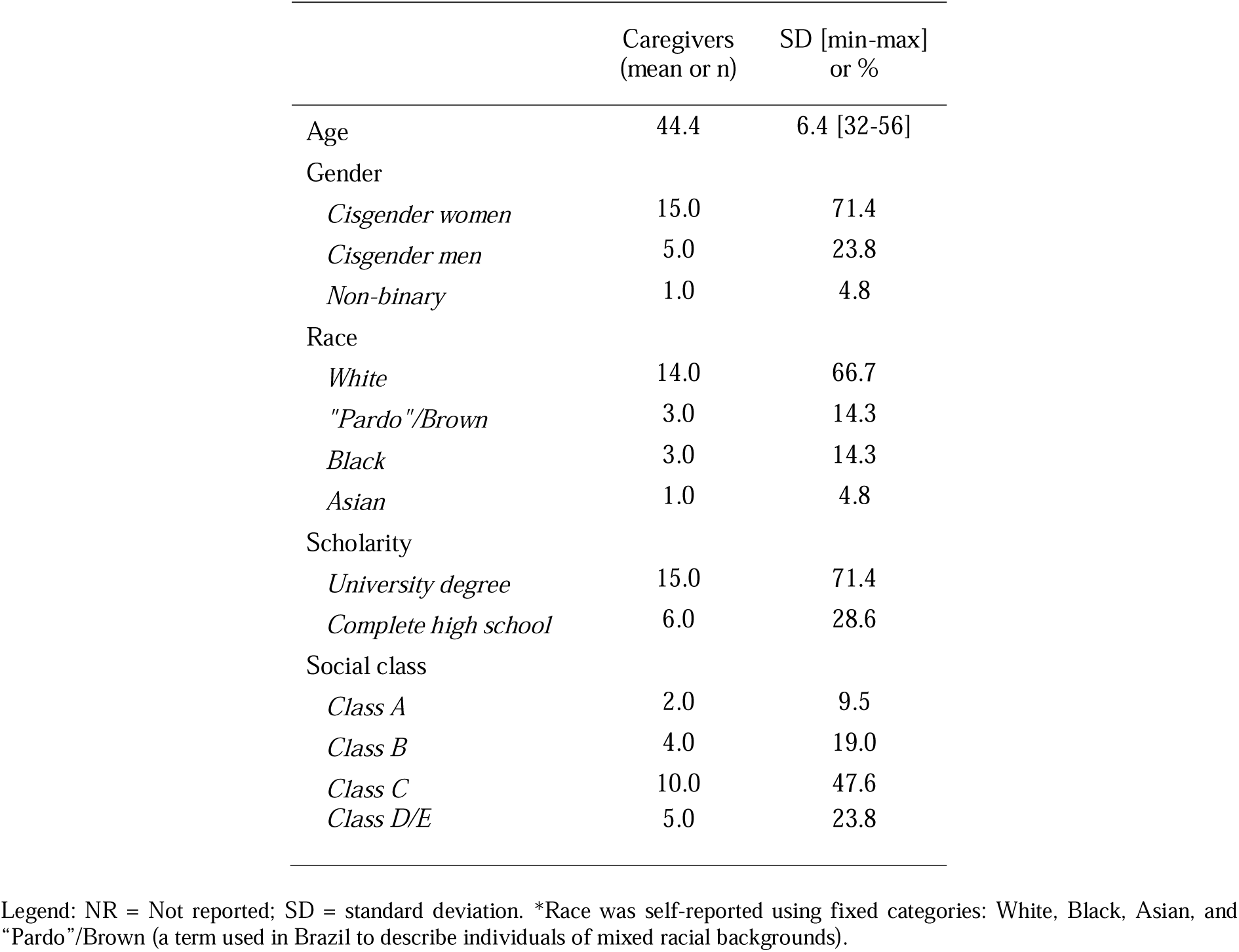
Demographic Characteristics of Caregivers (n=21).

|  | Caregivers<br>(mean or n) | SD [min-max]<br>or % |
| --- | --- | --- |
| Age | 44.4 | 6.4 [32-56] |
| Gender |  |  |
| *Cisgender women* | 15.0 | 71.4 |
| *Cisgender men* | 5.0 | 23.8 |
| *Non-binary* | 1.0 | 4.8 |
| Race |  |  |
| *White* | 14.0 | 66.7 |
| *"Pardo"/Brown* | 3.0 | 14.3 |
| *Black* | 3.0 | 14.3 |
| *Asian* | 1.0 | 4.8 |
| Scholarity |  |  |
| *University degree* | 15.0 | 71.4 |
| *Complete high school* | 6.0 | 28.6 |
| Social class |  |  |
| *Class A* | 2.0 | 9.5 |
| *Class B* | 4.0 | 19.0 |
| *Class C* | 10.0 | 47.6 |
| *Class D/E* | 5.0 | 23.8 |

Legend: NR = Not reported; SD = standard deviation. \*Race was self-reported using fixed categories: White, Black, Asian, and “Pardo”/Brown (a term used in Brazil to describe individuals of mixed racial backgrounds).

Transgender adolescents were mostly white (22 [71.0%]), from socioeconomic class C (13 [41.9%]), did not have any mental health condition (24 [77.4%]) and were going to school (30 [96.8%]) (Table 1). The majority of caregivers had university degrees (15 [71.4%]) and represented children and adolescents that were mostly white (14 [66.7%]), from socioeconomic class C (10 [47.6%]), that did not have any mental health condition (19 [90.5%]) and that were going to school (21 [100%]) (Table 1).

Of the 31 participating adolescents, only 1 reported using both male and female pronouns, all the remaining reported using pronouns according to the gender they identify with (i.e., female pronouns for trans girls and male pronouns for trans boys). Most participants aged 11 to 13 years considered themselves preteens and agreed that, depending on the situation, they act or prefer to act as child or adolescent. Those with ages between 14 and 18 considered themselves adolescents, with the exception of three participants, all aged >16, who declared themselves entering adulthood. Of the 21 participating caregivers, mostly were transgender children’s or adolescents’ parents, with the exception of an uncle and a grandmother. Only one identified as non-binary and used neutral pronouns, while all others were cisgender individuals reporting the use of pronouns in accordance with their gender identity.

The focus groups were conducted between 1 and 2 months after the CFM resolution was published. Transgender adolescents were aware of the new CFM resolution and, in some cases, familiar with the official justification presented, such as the alleged risk of “detransition.” However, they expressed uncertainty about practical details, including the minimum age for surgeries or whether those already on puberty suppression could begin hormone therapy at 16 years old. Conversations indicated that information circulated more through social media and peer-to-peer conversations than through clear institutional channels. Regarding caregivers, they were also aware of the new resolution, but presented varying levels of knowledge about it, with some admitting they were not fully up to date and trying to clarify questions throughout the group. Those who were more informed reported having followed discussions and changes from the beginning, through news, text messaging apps groups, and support networks. Some demonstrated partial understanding of the restrictions established by the resolution, particularly regarding their applicability to children and adolescents already undergoing therapies as stipulated by the previous resolution, for example, the continuation of puberty suppression or the transition to hormone therapy at age 16 for those already in puberty suppression.

The following key themes and subthemes were constructed: (a) Opinions and feelings about the new resolution - Opinions and feelings of transgender adolescents about the new resolution and Opinions and feelings of caregivers about the new resolution; (b) Conversation between child/adolescent and caregiver about the new resolution - Transgender adolescents’ view of the conversation and Caregivers’ view of the conversation; (c) Actions related to the new resolution; (d) Lived experiences opposing the new resolution; (e) Concerns regarding the consequences of the new resolution - Concerns of transgender adolescents about the new resolution and Concerns of caregivers about the new resolution (SM2).

### Opinions and feelings about the new resolution

#### Opinions and feelings of transgender adolescents about the new resolution

Transgender adolescents characterized the new resolution as “ridiculous”, “absurd”, and “wrong”, expressing feelings of despair, sadness, indignation, disappointment, and anger. Across narratives, they perceived the measure as an explicit expression of hate directed to them, framed more as an ideological than a health-based decision. As one participant argued:

> These are far-right government measures that are completely out of line and that aim only to take away people’s rights. But this, unfortunately, is nothing new […] Their intention isn’t to do something good for children, to protect children, to do something truly productive, it’s to hurt people, children, to increase rates of self-harm or suicide, for example. Anything they can do to take us away, they will do. (A29)

Adolescents emphasized that the core issue was not the gender-affirming therapies themselves - whether puberty blockers, hormone therapy, or surgeries - but rather the prejudice against transgender people’s existence and the denial of their autonomy in making decisions about their own lives. They expressed indignation at the idea that people with no transgender living experience were given authority over their futures.

Adolescents consistently framed this as a withdrawal of a right that is legitimately theirs. While some who had already accessed puberty blockers reported relief at not being directly affected, they nevertheless expressed distress on behalf of peers who would be deprived of such treatment:

> Anyway, on this issue *[puberty blocking]*, I still feel privileged, because there are trans people who will no longer have access to treatment. So, they will start developing the biological hormones they didn’t want. I, at least, have the possibility of being blocked. (A24)

A recurring opinion was the sense of injustice in the nonconformity between cisgender and transgender adolescents’ autonomy for changing their bodies. Participants stressed that cisgender children with precocious puberty continue to have access to blockers without restriction, while transgender ones are deprived of the same intervention. For those who had gone through puberty blocking, it was described as very beneficial, while for others, the new restrictions take away an option they considered essential to affirm their identity and decrease their gender dysphoria.

The postponement of the start of hormonal therapy until adulthood (previously allowed from age 16) was also met with discouragement. This was especially painful for those who had previously been unable to initiate hormones due to physical or mental health conditions, but who now, after a lot of time in follow-up, are ready to begin it. They emphasized that gender-affirming care is never granted hastily: it requires extensive clinical follow-up and a rigorous assessment process before access is authorized. Precisely because of this long monitoring, they argued that the risk of regret or detransition is drastically reduced. Thus, from their perspective, invoking detransition as justification for restricting access is invalid and dishonest. According to them, detransition often reflects external pressures - including religious, political, or familial coercion - rather than regret over gender identity (A27: “It could also be due to family issues. […] They say: ‘No, I am going to detransition to be accepted’. To have a house, to have a home.”).

They highlighted the inequality of denying transgender adolescents access to hormones and surgeries, so they can have bodies more congruent with their identity, based on age, while cisgender adolescents, often motivated by aesthetic concerns, are allowed to modify their bodies (e.g., cosmetic procedures) at a younger age, as long as their parents/caregivers agree.

> So, I found this decision completely ridiculous, because by making this law, they’re forcing children and teenagers to live in bodies that don’t belong to them. We don’t fit in with the body we were born with, and forcing us… (A26)

Surgical restrictions were less extensively discussed, however, they also reported being against the new resolution in this aspect. One participant noted the impracticality of access even under the new age limit of 21, recognizing the structural barriers that already made such procedures difficult to access.

### Opinions and feelings of caregivers about the new resolution

For caregivers, the resolution was perceived as a “nonsense” decision, an attack without scientific background, ideologically motivated, and a profound step backwards (C3: “I feel under attack, in a way, you know? […] and it caused me a lot of anxiety and distress”). They associated it with feelings of anger, sadness, despair, anxiety, anguish, indignation, and a sense of helplessness (C13: “You feel lost, right? Because it is like you are reaching a point, a bridge, and the bridge starts to collapse”).

For most, the resolution was understood as an ideological act, orchestrated by a largely conservative group of professionals within the medical field. They were critical of a medical class that often isolates itself, assumes superiority, and feels entitled to decide what is best for everyone. Some highlighted the contradiction of restricting gender-affirming care under the justification of potential health risks or regret, while allowing other procedures with equal or greater risks — such as cosmetic surgeries — to proceed without interference. In their view, the psychological impact of denying access to gender-affirming therapies for children and adolescents is far greater than the risks invoked as justification for prohibition. They argued that if health risks truly motivated these restrictions, attention should be directed toward procedures that affect broader populations, such as cosmetic surgery, rather than targeting a minority group.

The resolution was thus perceived as a direct attempt to prevent their child/adolescent from “being”, rather than simply limiting access to care (C8: “[*the possibility of continuing puberty blocking*] It’s being able to live. […] It’s being who you are. Without others pointing fingers”). They emphasized the injustice of strangers deciding what is best for their children, questioning the legitimacy of such authority:

> Who are you to decide, to say what is best for my child? Why do you have this power and then use it to do this? Why do you think you are doing them any good? It is horrible. You feel powerless at times. […] It is revolting. You have to wait for a human being to say ‘yes’ for you to see your child smile. (C8)

The restrictions were also seen as worsening inequalities, since only families with sufficient financial resources would be able to leave the country to undergo therapies abroad, prosecute the State or have access to therapies outside the Brazilian law (e.g., unregulated purchase of puberty blockers or hormones). Caregivers pointed out that transgender people live diverse realities that should be considered intersectionally - as they are not only transgender, but also white or black, rich or poor, thin or fat, with or without mental health conditions - and thus disproportionately vulnerable to compounded discrimination and barriers to care.

> When I started noticing people were talking about very high prices, I thought, and then I realized that transgenerity affects people transversally. There’s a class issue, like, present in this discussion. And then I said, guys, I think you don’t understand what we’re talking about. It’s not about thinking about yourself. We need to think about collective action, like, that really pays for everyone. […] And then, it’s also very interesting to see that, besides a given class issue, there’s also a racial issue. […] My son is black and trans. We have to discuss these things too. Intersectionality. (C7)

For parents whose children had already begun puberty blockers, they felt relief for their children not being affected, however, they expressed concern and sorrow for other families who would now be deprived of this opportunity. Puberty blocking was described as life-changing, offering children the chance to live and socialize without constant exposure to dysphoria, and to gain time for identity development without the distress of unwanted pubertal changes.

Moreover, caregivers fear for the children who were entering puberty at that time, recognizing the distress of knowing how much their children want to block puberty but not being able to support them in achieving it. They criticized the lack of scientific grounding for the decision, noting the contradiction in the simultaneous argument that more research is needed and then banning puberty blocking even in research contexts - which had been the only legal pathway until the resolution 2.427/2025:

> I don’t see any connection to science. We know all the care this outpatient clinic provides. And, in fact, there’s a resolution that is very contradictory. One thing, perhaps, you could say, for everything in Brazil, but in terms of research, we’re going to maintain it, because we must believe in something, we’re going to do research, right? And then it takes away the biggest reference centers. So, like, no more research, is that it? So, what do you want to base it on? Obviously, they don’t want to base it on anything, right? (C1)

Caregivers strongly rejected the argument that the risk of detransition justified the new restrictions, citing the lack of evidence that this occurs in a significant number of people. Many considered it unimaginable that their children, who had from an early age expressed themselves as transgender, would reverse this identity. Others took a more flexible stance, stating that even if their children decide to detransition, this would not represent regret but rather part of the natural process of self-discovery and reconstruction that characterizes human life.

> I draw parallels with other things. Until last year, my son did parkour. He loved it and went to the gym and all that. Suddenly, he started losing interest in parkour. He looked at other things, then he moved on to soccer, volleyball, and as a father, making this physical education comparison, I’m offering him all the opportunities and possibilities for him to see what he likes to do, what he wants. So he’s building, developing a taste for it, and changing it, and that’s okay. And the issue of identity is an additional layer, but we can draw a parallel with that. And we haven’t even touched on sexuality, which will be the same thing. So he’ll discover himself, get to know himself, experiment. And we’ll be there supporting him in these constructions and deconstructions. We are beings who are constantly deconstructing and reconstructing ourselves, and that’s why the question of regret doesn’t make sense. (C15)

Furthermore, they suggested that detransition was more likely among individuals without access to multidisciplinary professional support or adequate family acceptance. In their view, when such support exists, the chances of regret are minimal. For this reason, they advocated for expanding specialized clinics rather than restricting care.

Restrictions on hormone therapy and gender-affirming surgeries were not discussed as extensively, though most caregivers opposed them. Only two caregivers expressed reservations about starting hormone therapy at age 16, arguing that it required sufficient maturity and absence of confounding mental health conditions. Still, they emphasized that decisions about hormones and surgeries should be individualized, depending on multidisciplinary health team approval and each person’s readiness, and should not be curtailed by broad prohibitions.

### Conversations between transgender child/adolescent and caregivers about the new resolution Transgender adolescents’ view of the conversation

Transgender adolescents described the conversations with their caregivers about the new resolution as emotionally intense moments. Many highlighted the closeness with their mothers, who acted as trusted figures, both to explain and to provide comfort. Despite this support, the news was perceived as a form of loss and as a threat to rights already achieved. For all of them, caregivers acted as messengers and allies.

> My mom and I are very close […] she told me, ‘there’s now a new law that doesn’t allow blockers anymore.’ I said, ‘good thing I still have mine,’ but then she added, ‘when you’re 16 we don’t know if you’ll be able to start hormones.’ The ground fell from under me. But we talked, and she helped me. (A10)

### Caregivers’ view of the conversation

Caregivers of transgender children and adolescents described different strategies when addressing the new resolution with their sons and daughters. Children’s caregivers opted for protective silences, aiming to shield them from additional anxiety. This was particularly the case for those who were approaching puberty and anxiously awaiting the possibility of puberty blocking, as well as for younger children who were still far from this stage. In these cases, caregivers expressed concern that raising the issue prematurely could cause unnecessary distress. One caregiver reported deciding to remain silent and let the child bring up the topic if it emerged through other sources.

> I think it’s a very big thing for someone so small, with such a small heart, who longed and desired so much, because one of the tools for getting him here [*the outpatient clinic*] was the fact that he discovered and understood that there’s the possibility of blocking puberty and not growing a breast. So, for him, it’s the master. So, I didn’t say. And I also don’t know when, at what moment, right? I think if we’re already moved, imagine them, right? So, I chose not to say. (C6)

In contrast, most caregivers of adolescents chose a more straightforward discussion about the new resolution, sharing their indignation about the restrictions. One caregiver was the exception, who reported deliberately withholding the information from his son who was already receiving puberty blockers, reasoning that the resolution would not have any immediate effect on him and could cause unnecessary worry.

### Concerns related to the new resolution Concerns of transgender adolescents

The new resolution brought a wide range of concerns to the adolescents about their future. It is noteworthy that even those who had already done puberty blocking and/or started hormone therapy were worried by those who could not access them yet, demonstrating how they perceived the effects of the new resolution as a collective question. Adolescents were afraid that the restrictions lead transgender children to much more suffering, as it can be seen on the quote:

> Looking at myself, I feel relieved because I started much earlier, but at the same time, I look at the children who will come and I feel sad. And I also have…I’m almost certain that the rate of children self-harming will increase, so I feel even sadder, because I think some children just haven’t done it because of the [*puberty*] blocking. (A29)

Also, they were concerned about the possibility of transgender children and adolescents looking for illegal and/or insecure ways to get hormone therapy. Besides, most of the concerns were associated with the perspective of developing biological sex markers in discordance with their gender identity, which seems terrifying to them. It was possible to notice how anxious and distressed they were during the focus groups, imagining a future where they could not become who they are. Adolescents stated how they need and want to get access to puberty blocking, hormone therapy and gender affirming surgeries.

### Concerns of caregivers

Caregivers’ concerns about the future of their children and adolescents were especially related to the prejudice and violence that they may experience without getting access to gender affirming therapies. They consider that by living in a society that is transphobic, transgender people will always have to face these prejudices in their lives, which could be reduced by accessing these therapies. One of them stated:

> I get worried too, like, without [*puberty*] blocking, others pointing it out. If you can’t get a job, you face prejudice. That’s why so many people end up marginalized and unemployed, right? You can see so many trans people unemployed, right? Because they’re marginalized, there’s no way around it. It’s hypocritical to say otherwise. Appearances, unfortunately, count for a lot, right? Appearances count for a lot. (C13)

Similar to adolescents, caregivers were worried about the people who will not be able to access adequate care for their own children. Both caregivers and adolescents demonstrated a strong sense of community, with some caregivers addressing socioeconomic and racial inequalities that may restrict possibilities. They also alleged being worried with a probable search by transgender people for illegal and unsafe ways to get hormones and relatable strategies to modify their bodies.

It is interesting to notice that many of them reported having conversations with their children about how they must love and have a good relationship with their bodies, while waiting to be able to access gender affirming therapies. Caregivers stated that transgender children and adolescents having their puberty blocked is a way to guarantee that they will have time to get more mature and understand who they are and what they want from their bodies.

### Lived experiences opposing the new resolution

All of the transgender children and adolescents who had already started puberty blocking and hormone therapy stated that it was very important and positive to them. Especially considering that the gender affirming therapies were offered at the specialized outpatient gender identity service, they demonstrated being confident about the safety of their effects. There were no reported physical, emotional or social disadvantages. Both strategies were mentioned in different ways: by blocking their puberty, adolescents were satisfied with not developing biological body changes of their sex assigned at birth, such as breasts, body hair and tone of voice (A19: “[*puberty*] Blocking is really good. Like, everything that I was afraid to change at the beginning of transition, with [*puberty*] blocking, they didn’t. So, it makes me feel much calmer”); with hormone therapy, they were happy with the development of physical characteristics that match with what is expected for their gender identity, which was their wish when they started treatment:

> My waist is much more defined and feminine than it was when I only used puberty blockers, right? So, my waist movement, I think it shifted a lot of my fat here to my butt and such, and my breasts grew. They didn’t grow that much, but they grew a little. So, it’s something that I feel is changing (…) that’s what I thought the hormones would do to me and I am very happy. (A29)

They always demonstrated great certainty and clarity about how they want their bodies to look like, and being able to achieve this reassured them. The absence of gender affirming therapies - whether imagined as a possibility or experienced as a reality - was associated with intense psychological and social suffering, captured in its most extreme form by the following quote:

> A22: It’s like, I think if I hadn’t [puberty] blocking, I would want to kill myself. A24: It’s true.
>
> LCO: And speaking of hormone therapy, surgery, what do you think about these things? Hormonal therapy, surgery… Do you think you need to go through these things?
>
> A21: I think I would be happier.
>
> LCO: Would you be happier?
>
> A21: Yes, I think I need to go through this. LCO: All these things?
>
> A21: Yes.

### Actions related to the new resolution

There were different points of view and perceptions about what has been done to confront the new resolution, and some direct actions against it. In general, initially, both caregivers and transgender adolescents were surprised and took a while to understand what was at stake. Even so, most caregivers quickly organized collective actions aimed to legally confront CFM, compounding crowdfunding, petitions and protests. It is noteworthy that even those caregivers of children who, in principle, would not be affected by the new resolution, joined the actions with those who were immediately affected.

Besides recognizing the efforts of their caregivers, transgender adolescents were disappointed with the omission of the major society, especially of those who are part of the LGBTQIAPN+ community. They mentioned that they have not seen any famous transgender person talking about the new resolution. The São Paulo LGBTQIAPN+ Parade was about to take place around the same time this research took place, and adolescents pointed out how this celebration was, in its roots, a day of fighting for community rights, and how nowadays they perceive it as just a party, which often makes the fights of the transgender population invisible.

Both caregivers and transgender adolescents stated that participating in this research was also a way to act against the new resolution, as one of the most important arguments used by CFM was the scarcity of researches that guarantee the safety of gender affirming therapies. Finally, caregivers mentioned how the imposition of these restrictions made them reflect about their political and social compromises with transgender rights. Even if they have always been compromised with their children, now they perceived how this commitment cannot be restricted to the private sphere, as exemplified by the quote:

And the way we’re organizing to take action, perhaps, is that everyone is here. The story of the Minha Criança Trans [*“My Trans Child”, a Non-Governmental Organization*] movement, we’re joining in, we’re participating in these paths that we believe are important. And another thing is that we felt we needed to be more active. So, until then, we lived here—in fact, we lived and still live in our own world here, dealing with [*children’s name*] issues individually. And we started to think we needed to be more active, and be present in places, and really be there. We might not be able to do much concretely, but we need to be there. So we started going to the acts. We went to the Legislative Assembly here in São Paulo, where there was a hearing. [*children’s name*] is here, waiting for her mother to arrive to pick her up. And we’re organizing to go to the [*LGBT Pride*] parade, to participate. Anyway, we’re in this movement of feeling that we have to be in places. Apply pressure, participate, beyond this individual thing that we already take care of and have always taken care of, because we need something more collective. That’s how things hit us, in the early moments, post- news, etc. (C3)

## DISCUSSION

The findings of this study show how the CFM new resolution was experienced by transgender adolescents and their caregivers, as well as caregivers of children as an act of political persecution and a deliberate attempt to deny rights, rather than a neutral regulatory adjustment grounded in evidence-based medicine. This perception resonates with those from researchers and clinicians working with this population, which have identified how restrictions on gender-affirming care are increasingly deployed as tools of ideological contestation rather than medical prudence (Billard, 2024; Longobardi et al., 2025).

For transgender adolescents, the resolution carried existential weight. Their narratives were filled with despair, sadness, anger, and indignation, but also with critiques of the contradictions underpinning the decision. They emphasized that cisgender children continue to access puberty blockers to treat precocious puberty, while transgender ones are being denied the same intervention for gender dysphoria. Similarly, they noted that cisgender adolescents face no barriers when seeking elective procedures such as cosmetic surgery, while transgender ones are deprived of care that is essential to their mental health, well-being, and identity. In line with prior studies, these accounts underscore how access to gender-affirming interventions is not about preference or aesthetics, but about aligning one’s body with one’s identity in ways that improve mental health and quality of life that can be life-saving (Costa et al., 2015; de Vries et al., 2014; de Vries et al., 2011; Kuper et al., 2020; Kristina R. Olson, 2025; Tordoff et al., 2022; Turban et al., 2020).

A central theme across the groups was the invalidity of detransition as a justification for restricting care. Both adolescents and caregivers pointed out that access to any gender-affirming intervention is preceded by long-term and multidisciplinary clinical follow-up, under robust protocols implemented in outpatient clinics affiliated with university hospitals, which was in line with the 2019 CFM resolution (Medicina, 2019). This process was described as exhaustive but necessary, mainly because it reduces the likelihood of regret. Adolescents were emphatic that invoking detransition ignores the reality and often misrepresents the phenomenon: in their view, detransition frequently reflects external pressures such as family rejection or religious coercion, rather than regret itself. Caregivers added that when professional and familial support are available, as in the specialized clinic setting, the risk of regret becomes vanishingly small. These insights are in line with evidence demonstrating that regret following gender-affirming care is rare (Cavve et al., 2024; K. R. Olson et al., 2024; Turban et al., 2020; van der Loos et al., 2022) and that outcomes are overwhelmingly positive when care is provided according to established standards (Bränström & Pachankis, 2020; Coleman et al., 2022). Likewise, at our clinic, where the previous CFM resolution guided care for nearly a decade, 79 transgender adolescents were treated with puberty blockers, and only one later chose to detransition, without regret.

The postponement of access to hormones until the age of 18 was perceived as especially unfair. Adolescents who underwent years of monitoring and were finally reaching a stage of viability for treatment initiation now found themselves prohibited to follow through with hormone therapy. Caregivers also reported this sense of injustice, with some highlighting that puberty blocking and hormone therapy can be decisive in allowing children and adolescents to live, socialize, and flourish without the constant distress of gender dysphoria and transphobia. The minority of caregivers expressed caution about initiating hormones at 16, citing concerns about maturity in the context of specific mental health conditions; however, even them emphasized that such decisions should remain individualized rather than determined by broad prohibitions. Surgical restrictions were less discussed, nonetheless, both adolescents and caregivers interpreted the new age threshold as another manifestation of systemic denial. As WPATH and the Endocrine Society emphasize, puberty blocking effects are fully reversible once use is discontinued. Furthermore, hormone therapy and surgical interventions are always preceded by rigorous evaluation, with surgeries typically being considered only for late adolescents or adults (Coleman et al., 2022; Hembree et al., 2017), making the blanket restriction appear less about medical caution and more about symbolic exclusion.

The conversations within families about the resolution also shed light on strategies of care and resistance. Some caregivers, especially of younger children, chose protective silences believing that disclosing the restrictions prematurely would overwhelm them with anxiety. Others opted to openly discuss the resolution and share their indignation as a way to validate their children’s experiences. These different strategies illustrate how caregivers attempt to mediate between the structural violence of policy and the everyday emotional needs of their children. Research on parental support for transgender adolescents consistently demonstrates that such familial support is one of the strongest protective factors against poor mental health outcomes (Simons et al., 2013).

At the same time, both adolescents and caregivers recognized the broader structural consequences of the resolution. Families noted that only those with financial resources would be able to circumvent restrictions through legal action or private alternatives, while poorer and more marginalized families would be left with no options. This awareness reflects an intersectional perspective, recognizing that transgender adolescents are not only transgender but also differently positioned across race, class, and health status - and that these intersections shape their vulnerability to exclusion. The concern that inequalities will be amplified by the new resolution aligns with findings that restrictions on gender-affirming care disproportionately harm those who are already marginalized (Puckett et al., 2019).

Finally, while despair and anger were recurrent, there were also accounts of mobilization. Caregivers reported resistance through involvement in collective actions, such as petitions, lawsuits, and protests, and creation of support nets (which is also known as “*aquilombamento”*) as collective strategies of survival and political agency (Aranha et al., 2023). In contrast, transgender youth expressed frustration with what they perceived as a lack of visible support from broader LGBTQIAPN+ movements. This perception diverges from reports that emphasize the centrality of this community in defending their rights, visibility, and promoting equality (Aranha et al., 2023). This divergence highlights adolescents’ perception that those at the forefront of the fight against the new resolution are their closest support networks, such as their caregivers, health professionals from their outpatient clinic, and non-governmental organizations directly involved in guaranteeing the rights of transgender people. Furthermore, it highlights the need to understand how forms of resistance are unevenly distributed and perceived among different members of the “same” community and how these gaps can shape both political engagement and the production of health and belonging.

The main strength of this study is its timely and rigorous qualitative assessment of transgender people and their caregivers at the largest specialized clinic in Latin America. By amplifying the voices of this vulnerable group in the face of a resolution that directly affects them, the study informs the medical and public debate on the injustice and disrespect for human rights, especially regarding the ontological right to existence of the new measures, about “being who you are” and feeling “able to live”. Moreover, participants demonstrated a strong and accurate understanding of the resolution’s content and its practical implications during the focus groups. Limitations include the single-center design, possible selection bias given that only those who agreed to participate were assessed, and the possibility that some participants had incomplete knowledge of the resolution, which may have influenced their perceptions (e.g., those who mistakenly believed the resolution would close the clinic might have been more resistant to it). In addition, our findings and conclusions are based on a sample of adolescents who had access to a multidisciplinary treatment service, who were mostly white and from middle-class backgrounds (social classes B and C). This profile may not broadly reflect the experiences of transgender adolescents in Brazil, particularly those from racialized groups, lower-income families, or without access to specialized care. Further research, especially with an explicitly intersectional perspective, is needed to capture this diversity of realities. Nevertheless, as a qualitative study, our aim was not generalization, but rather an in-depth exploration of meanings and experiences.

Taken together, findings show that the resolution was not experienced merely as a change in medical regulation, but as a profound assault on rights, health, and dignity of transgender children and adolescents. By integrating their lived experiences with critiques of scientific misrepresentation, adolescents and caregivers illuminate the contradictions and harms of restrictive policies. In doing so, they emphasize that health policies should be informed not only by international research and clinical experience, but most crucially, must be validated and contextualized by national studies. These studies must consider the reality of transgender individuals and the country’s health system, as well as the clinical protocols established in reference centers for their care. Beyond clinical implications, the narratives also reveal how such restrictive policies exacerbate stigma and exclusion while simultaneously mobilizing families and adolescents toward collective resistance. Evidence-based treatments and policies should not leave out transgender voices.

## Data Availability

All data produced in the present study are available upon reasonable request to the authors.

## Author Contributions

All authors had full access to all of the data in the study and took responsibility for the integrity of the data and the accuracy of the data analysis.

Concept and design: Gualano, Saadeh, Roschel, Scagliusi, Mazzolani.

Acquisition, analysis, or interpretation of data: Mazzolani, Longobardi, Smaira, Sabatini, Evangelista, Oliveira, Torres, Bittar.

Critical review of the manuscript for important intellectual content: All authors. Administrative, technical, or material support: Saadeh, Scagliusi.

Supervision: Gualano, Saadeh, Roschel, Scagliusi.

## Data Sharing Statement

The data that support the findings of this study are available from the corresponding author upon reasonable request.

