## Supplementary Material for "“I feel under attack”: Transgender Adolescents and Caregivers Opposition to Restrictions on Gender-Affirming Medical Care in Brazil"

**Supplementary materials**

**SM 1. Semi-structured interview guide**

**TRANSGENDER ADOLESCENTS'S VERSION**

Welcome and explanation of the meeting: "Today we're going to chat about family, school, physical activity, eating, how we feel about our bodies, puberty blocking, hormone therapy..."

Explanation of how the group works: "There are no right or wrong answers; we want to know what you think."

Icebreaker: Each child shares their name, pronouns and a physical activity or food they really enjoy.

**Family**

1. Let's start by talking about your home and family... What is your house/apartment like? Who lives with you? (*probes to understand who the parents are, if you're an only child, how many people live in the house and interact with this teenager*) What do you usually do together? (*probes to understand the relationship with family members, whether they do more sedentary or active activities, socioeconomic profile depending on what they have available at home to do*)

**Childhood, Adolescence, and Relationship with the Body**

1. Now tell me: do you consider yourselves children or adolescents? And what's the difference between being a child and being an adolescent?
2. And how do you spend your free time? Who do you like to spend time with? And when you're together, what do you like to do?
3. I wanted to talk about bodies, yours and others'. How do you feel about your own bodies? Have you ever compared or do you compare yours to other people's bodies? Who are these people? (*probes to understand whether they are cis or trans, famous or well-known*) Have you ever done anything to change the appearance of your bodies? How was it? (*probes to understand if it was before or after transition, if you suffered from comments about your bodies*)
4. Now, can you share with me what it is like for you to think about going through puberty or what it was like to go through puberty?

**School**

1. Let's now talk about your school. Where do you study (*probes to understand if it's public or private*)? What do you think of your schools? (*probes to understand if it's an environment where adolescents face any prejudice, if they feel like they belong*)
2. Do you eat at your schools? If so, what is it like to eat at school? (*probes to understand what they eat, if they bring anything, if they buy it, who prepares it*)
3. What's the food like at your house? (*probes to understand what they eat, with whom, where, who's responsible, who cooks*) Do you think the food at your house is healthy (*probes to explore the reasons and what could be different*)? What about the food at school? Still speaking of school, when do you move your body there? (*probes to explore break time, physical education class, and extracurricular activities*) And what do you think of physical education class? (*probes to understand if bullying happens, if the teacher divides the class between boys and girls, difficulties related to clothing*)
4. How would you help a friend be healthier? And how would you help them feel better about their own body?

**Gender-affirming therapies**

1. And going to the doctor, what is it like for you? How do you feel? (*probes to understand prejudice in these settings, to understand if there is a difference between those in the outpatient clinic and those outside*)
2. What would it be like to be able to block puberty, or what is it like to be able to block puberty? And what is it like to think about hormone therapy and gender-affirming surgeries? Do you feel you need to go through them? What do you hope will happen if you go through them?
3. Did you hear about the new resolution from the Federal Council of Medicine? (*If not, explain*) How do you feel about the news that puberty blocking may no longer be possible? What do you think of the justification for this decision? (*State the reasoning if they don't know*)
4. And how do you feel about the news that the minimum age for hormone therapy may become 18 and for some gender-affirming surgeries 21? What do you think of the justification for this decision? (*State the reasoning if they don't know*)
5. What have you and your guardians been saying about this news? What have you been doing or would like to do about it?

**Future**

1. What are your plans for when you become adults?
2. What should the world/society be like for you to have a healthier and happier life?

**CAREGIVERS’ VERSION**

Welcome and explanation of the meeting: "Today we're going to chat about you and your children/adolescents. We'll talk about family, eating, their school, activities they enjoy, their relationship with their bodies, questions about puberty blocking, hormone therapy..."

Explanation of how the group works: "There are no right or wrong answers; we want to know what you think."

Icebreaker: Each caregiver shares their name and pronouns, the name of the child/teen they're caring for, the child/teen's age, their relationship, and an activity they enjoy doing or a food they enjoy sharing with their child.

**Family**

Trigger question: "Let's start by talking about your home and family... What's your home and your daily life like as a family?"

Probes (if necessary):

- Who lives in the house with you?
- Do you have other children/adolescents besides your transgender children/adolescents?
- What do you usually do together at home? (More active or sedentary activities?)
- What is your relationship with your transgender child/adolescents? And what about that of other family members?
- Do you have contact with other transgender people (adults or children)? Is this contact inside or outside the outpatient clinic for transgender adolescents?

**Childhood, Adolescence, and Relationship with the Body**

Trigger question: "Do you consider your children a children or a teenager? Why? What has this phase been like?"

Probes (if necessary):

- What do they usually do in their free time?
- Who do they like to spend this time with?
- How do you perceive their relationship with their own bodies?
- Do they often compare themselves to other people? With whom? (cis or trans, famous, well-known...)
- Have you ever done anything to change your appearance? What was it like?
- How do you feel about your own bodies?
- What is it like for you to think about your children's puberty (with or without blockers)?
- How do you perceive them feeling about puberty?

**School**

Trigger question: "Let's now talk about the school your children/adolescents attend. Where do they study? What do you think of your children's schools?"

Probes (if necessary):

- Where do they study? Is it public or private?
- Have they experienced prejudice or problems at school?
- Trigger question: "Do they eat at school? What do they say about it?"
- Do they bring food, buy it, and who prepares it?
- And at home, what is the diet like? What do they usually eat, where, and with whom?
- Who cooks? Is the food at home considered healthy?
- Trigger question: "At school, when do they move around (recess, physical education, extracurricular activities)?"
- What do they say about physical education class?
- Do they like it?
- Do they get bullied?
- Is the class divided into boys and girls?
- Do they feel uncomfortable with their clothing?

Trigger question: "What do you think would help your children/adolescents be healthier?"

Probes (if necessary):

- And what would help them feel better about their bodies?

**Gender-affirming therapies**

Trigger question: "And what is it like for you to take your children/adolescents to the doctor? How do you and your children/adolescents feel?"

Probes (if necessary):

- Have you ever experienced prejudice in any service? Is there a difference between transgender outpatient clinic care at and outside it?

Trigger question: "Did you hear about the new CFM resolution? How did you feel about this news?"

Probes (if necessary):

- What did you think of the justifications given for this decision?
- Have you told your children about this change? How have you talked about it at home?
- Have you done or thought about doing anything in response to this resolution?

Trigger question: Can you share with us what it's like for you to think about your children/adolescents undergoing puberty blocking, hormone therapy, and gender-affirming surgeries?

- Puberty blocking: How do you think they feel about this possibility?
- Hormone therapy and gender-affirming surgeries: Do you think they want or need to go through these processes?
- What do you think they expect to happen after undergoing these procedures?

**Future**

Trigger question: "When you think about your children's/adolescents's future, what do you imagine?"

Probes (if necessary):

- What do you imagine they will be like in a few years?
- What are your dreams for them?
- And do you know what their dreams are?
- What do you think would help make these dreams come true?
- What should the world/society be like so that you and your children can have healthier and happier lives?
- What still needs to change?

**SM 2. Codebook**

| **Code** | | **Short name** | **Short description** | **Detailed description** | **Inclusion criteria** | **Exclusion criteria** | **Typical example** | **Atypical example** | **Close but no** |
| --- | --- | --- | --- | --- | --- | --- | --- | --- | --- |
| **Theme** | **Subtheme** |  |  |  |  |  |  |  |  |
| Opinions and feelings about the new resolution | Opinions and feelings of transgender adolescents about the new resolution | Opinions and feelings of transgender adolescents | What transgender adolescents think and how they feel about Brazil’s Federal Council of Medicine resolution (2.427/2025) restrictions on gender-affirming care. | The central focus of this theme is adolescents’ opinions and feelings about the new resolution. We aim to understand what they think about and which are their emotional responses to the imposed restrictions, the justifications used, the medical community, the actions/movements resulting from the resolution, and its consequences for their lives and the lives of other trans people. | Includes statements in which adolescents express opposing or neutral opinions, sensations or emotions related to the restrictions imposed by the new resolution, particularly concerning puberty blockers, hormone therapy, and gender-affirming surgeries. It also includes opinions, sensations or emotions related to the justifications for the resolution, the medical community, and the personal, collective, and legal actions/movements arising from the resolution and its consequences for their lives and the lives of other trans people. | It does not include trangender adolescents's opinions, sensations or emotions related to other subjects as well as their actions related to the new resolution. | I think it's ignorant on their part, especially because the CFM is there as professionals to serve the Brazilian people as a whole. So when they're taking away our rights and they don't show any proven data that it's harmful in any way, they're putting their own interests above ours, and I think that's ridiculous. It's disrupting so many lives. And like, for all the trans people who are undergoing treatment, I feel terrible for myself too, because I believe that, for the vast majority, it's a dream to go through a safe transition, conscious of what they're doing, and they're getting in the way of that. (Y30) | And then it started blocking puberty really well, but then this new law came along. And then, right in this moment, my doctor at the Children's Institute said I had to increase my dose. But I can't. Because of the law. So, wow, I started crying at the Children's Institute, but then I got home with my mom and she said, "Why are you crying? If they don't block you, I will." But she won't do that because she knows the risks. She was just joking to make me feel better, because I was really sad. (Y10) | As someone who went through this hormonal transition, I feel like it helped me a lot, it did a lot for me, it did a lot for my self-esteem, for my image, it helped me with my dysphoria, which is a very, very real problem. (Y29) |
|  | Opinions and feelings of caregivers about the new resolution | Opinions and feelings of caregivers | What transgender children and adolescents caregivers think and how they feel about Brazil’s Federal Council of Medicine resolution (2.427/2025) restrictions on gender-affirming care. | The central focus of this theme is the caregivers’ opinions and feelings about the new resolution. We aim to understand what they think about and which are their emotional responses to the imposed restrictions, the justifications used, the medical community, the actions/movements resulting from the resolution, and its consequences for the lives of those they care for and other trans people. | Includes statements in which caregivers express opposing, neutral, or favorable opinions, sensations or emotions related to the restrictions imposed by the new resolution, particularly concerning puberty blockers, hormone therapy, and gender-affirming surgeries. It also includes opinions, sensations or emotions related to the justifications for the resolution, the medical community, and the personal, collective, and legal actions/movements arising from the resolution and its consequences for those they care for and other trans people. | It does not include caregiver's opinions, sensations or emotions related to other subjects as well as their actions related to the new resolution. | My opinion about the resolution you asked about is also, it is complete nonsense, it has no scientific basis, it is really a political persecution. (C3) | I'm not in favor of hormonization at 16, personally, folks. And I think it depends on a number of situations, it depends on... it's very specific to each person. In my case, I'm not in favor, especially with all the disorders he already has. I think this is a very personal issue. (C21) |  |
| Actions related to the new resolution | | Actions | What transgender adolescents, caregivers and other individuals or communities are doing about Brazil’s Federal Council of Medicine resolution (2.427/2025) restrictions on gender-affirming care. | This theme focuses on what adolescents and caregivers are doing and their perceptions about what others are doing in response to the new resolution. The aim is to understand the actions and behaviors they have taken to confront or cope with the imposed restrictions, the justifications used, the medical community, and the consequences of the resolution for the lives of trans people. | Includes statements in which adolescents or caregivers describe theirs and others' people/groups actions, whether positive, neutral, or negative, in relation to the restrictions imposed by the new resolution—particularly regarding puberty blockers, hormone therapy, and gender-affirming surgeries—as well as actions related to the medical community, and personal, collective, and legal responses to the resolution and its consequences for the lives of trans people. | It does not include actions related to other subjects as well as opinions and feelings related to the imposed restrictions. | And we kept mobilized within the group. She mentioned the group that was coming here. So, we organized a major mobilization. The group contacted the law firm to file a lawsuit for those who were here at the outpatient clinic about to begin the [puberty] blocking. We were willing to split the costs with the lawyer and everything. Our contribution, even if it doesn't impact [children's name] at this moment, but we continue to mobilize collectively to overturn, to contribute to fighting this decision which, for us, was clearly, as I said, a political decision, with a shallow basis of research that doesn't support it.  These are research from abroad that doesn't justify it, anyway. We, as I said, are fighting and hoping that it will be overturned, finally, that it can, it will be reviewed. (C15) | But that's it, if I can give my opinion, talk about it, protest, I will protest, I will speak out against it, I will be against it, I will be the most outrageous opposition this country has ever seen. But that's it, I'm against it, and wherever and however I can be against it, I will be against it. (Y29) |  |
| Concerns regarding the consequences of the new resolution | Concerns of transgender adolescents about the new resolution | Concerns of transgender adolescents | What transgender adolescents are worried will happen as a consequence of Brazil’s Federal Council of Medicine resolution (2.427/2025) restrictions on gender-affirming care. | The essence of this theme is to capture what adolescents think might or will happen as a result of the new resolution imposing restrictions on gender-affirming therapies. The aim is to understand the projections they make regarding health, quality of life, and social (and political) integration in a scenario where the resolution is enforced. | Includes statements in which adolescents describe how they believe the restrictions imposed by the resolution will affect their lives (physical and mental health, social interactions), the lives of other trans people, and the professionals involved in trans care, as well as society in general. | It does not include trangender adolescents'concerns related to other subjects or current emotions or actions already taken. | "And I also have...  I'm almost certain that the rate of children self-harming will increase, so I feel even sadder, because I think some children just haven't done it because of the [puberty] blocking, because most trans children have this dysphoria and the [puberty] blocking causes less dysphoria, so it will take away the only thing that makes these children happy." (Y28) |  |  |
|  | Concerns of caregivers about the new resolution | Concerns of caregivers | What transgender children and adolescents caregivers are worried will happen as a consequence of Brazil’s Federal Council of Medicine resolution (2.427/2025) restrictions on gender-affirming care. | The essence of this theme is to capture what caregivers think might or will happen as a result of the new resolution imposing restrictions on gender-affirming therapies. The aim is to understand the projections they make regarding health, quality of life, and social (and political) integration in a scenario where the resolution is enforced. | Includes statements in which caregivers describe how they believe the restrictions imposed by the resolution will affect the lives of those they care for (physical and mental health, social interactions), the lives of other trans people, and the professionals involved in trans care, as well as society in general. | It does not include caregiver's concerns related to other subjects or current emotions or actions already taken. | I'm also worried, like, without [puberty] blocking, others pointing it out. You can't get a job, because you'll just keep a job, people face prejudice. That's why so many people end up marginalized and unemployed, right? You can see so many trans people unemployed, right? Because they're marginalized, there's no way around it. It's hypocritical to say otherwise  Looks, unfortunately, count for a lot, right? Looks count for a lot. (C13) |  |  |
| Conversation between transgender child/adolescent and caregiver about the new resolution | Transgender adolescents' view of the conversation | Transgender adolescents' view | What was the debate like and what arose between the adolescents and their caregivers regarding the resolution from the adolescents's perspective. | This theme explores what adolescents say and think about having conversations with their caregivers regarding the new resolution. We are interested in whether such conversations occurred from the adolescents`s point of view, as well as their feelings and opinions about the conversation. | Includes statements in which adolescents describe whether their caregivers talked to them about the new resolution, what was shared in that conversation, how they felt about it, whether they found it important to have such a conversation, or whether they would like to have it if it hasn’t occurred yet. | It does not include transgender adolescents's view of conversation with other people or about other subjects. | Then I said, "Wow, but I didn't know, what happened?". She explained it to me, then my parents showed their indignation, and that was basically it, like, we talked about it. (Y28) | But when she told me, I was very upset. But she said everything would be okay, right? It's not His will, I mean... She told me that everything is in God's time, if God wants it that way... It's because it has to be that way, then I felt more at ease. (Y31) |  |
|  | Caregivers' view of the conversation | Caregivers' view | What was the debate like and what arose between the adolescents and their caregivers regarding the resolution from the caregiver's perspective. | This theme explores what caregivers say and think about having conversations with children/adolescents regarding the new resolution. We are interested in whether such conversations occurred from the caregivers’ point of view, as well as their feelings and opinions about the conversation. | Includes statements in which caregivers describe whether their children/adolescents talked to them about the new resolution, what was shared in that conversation, how they felt about it, whether they found it important to have such a conversation, or whether they would like to have it if it hasn’t occurred yet. | It does not include caregivers' view of conversation with other people or about other subjects. | We decided not to share it with her, we talked about it, about this possibility, and we decided not to do it, because we thought it wouldn't be something either, it wouldn't bring anything, this super-sincerity wouldn't help at all. (C3) | Regarding my son, as I said, he doesn't yet have that dimension of activism, so we don't... He didn't know, we didn't talk about it, there's no... It didn't have an impact on him, in that sense. (C15) |  |
| Lived experiences opposing the new resolution | | Experiences | What were the experiences that transgender adolescents and caregivers' report that contradict the new resolution. | This theme focuses on what adolescents and caregivers report about their own experiences or those of others that contradict or challenge the new resolution. We aim to understand whether they had such experiences themselves or heard about them from others, and how these experiences relate to arguments against the restrictions and justifications imposed by the resolution. | Statements from transgender adolescents and caregivers about their personal experiences, those of people they know, or others, that contradict or challenge the assumptions or restrictions imposed by the new resolution (e.g., positively impact their or others' health, well-being, and identity). | It does not include transgender adolescents or caregivers' statements of their experiences or from others, related to gender-affirming care that were not directly associated with its impact on the new resolution or any experience related to other subjects. | "So,[puberty] blocking is really good. Like, everything that I was afraid of at the beginning of the transition, with the blocking, didn't. So, like, it makes me feel much calmer." (Y19) |  |  |
